# Clinical Characteristics, Risk Factors and Outcomes Among the First Consecutive 1,096 Patients Diagnosed with COVID-19: The Kuwait Experience

**DOI:** 10.1101/2020.05.09.20096495

**Authors:** Sulaiman Almazeedi, Sarah Al-Youha, Mohammad H. Jamal, Mohannad Al-Haddad, Ali Al-Muhaini, Fahad Al-Ghimlas, Salman Al-Sabah

## Abstract

**Background:** In Kuwait, prior to the first case of COVID-19 being reported in the country, mass screening of incoming travelers from countries with known outbreaks was performed and resulted in the first identified cases in the country. All COVID-19 cases at the time and subsequently after, were transferred to a single center, Jaber Al-Ahmad Al-Sabah Hospital, where the patients received standardized investigations and treatments. The objective of this study was to characterize the demographics, clinical manifestations and outcomes in this unique patient population.

**Methods:** This retrospective cohort study was conducted between 24^th^ February 2020 and 20^th^ April 2020. All consecutive patients in the entire State of Kuwait diagnosed with COVID-19 according to WHO guidelines and admitted to Jaber Al-Ahmad Al-Sabah Hospital were recruited. Patients received standardized investigations and treatments. Multivariable analysis was used to determine the associations between risk factors and outcomes.

**Findings:** Of 1096 patients, the median age was 41 years and 81% of patients were male. Most patients were asymptomatic on admission (49.5%), 69.4% had no signs of infection and 94.6% were afebrile. Only 3.6% of patients required an ICU admission and 1.7% were dead at the study’s cutoff date. On multivariate analysis, the risk factors found to be significantly associated with admission to intensive care were age above 50 years old, a qSOFA score above 0, smoking, elevated CRP and elevated procalcitonin levels. Asthma, smoking and elevated procalcitonin levels correlated significantly with mortality in our cohort.To our knowledge, this is the first large retrospective cohort study observing the characteristics of the initial consecutive COVID-19 patients of an entire country. Further, large proportion of asymptomatic patients provides novel insights into the clinical features of patients with milder disease.

**Funding:** Research Grant Awarded by the Kuwait Foundation for the Advancement of Science.

## Introduction

Defining the clinical characteristics and associated outcomes of patients diagnosed with coronavirus disease (COVID-19) is integral to improving our understanding and management of this disease. Several articles have recently been published, describing the clinical features and outcomes of retrospective cohorts of patients with COVID-19^1–3^ Most of the patients included in those studies were deemed sufficiently ill to merit being hospitalized. As a result, the clinical features and outcomes that have been described are representative of those symptomatic patients.

The COVID-19 patient cohort in the State of Kuwait is unique for several reasons. As soon as reports emerged of an outbreak in certain regions in Iran, all citizens were immediately repatriated, and mass screening of all travelers for COVID-19 was implemented. All the cases, irrespective of symptoms, that tested positive for SARS-COV-2 were hospitalized and remained hospitalized until two negative polymerase chain reaction (PCR) results were obtained from nasopharyngeal swabs. As the disease became more widespread, the same protocol was implemented for COVID-19 cases from non-travelers. Consequently, all patients diagnosed with COVID-19, early on Kuwait, received standardized investigations and treatments in the same treating facility. This presents an opportunity to obtain a more holistic understanding of the clinical features and outcomes of patients diagnosed with COVID-19, including patients who present with no symptoms. The objective of this study is to summarize the clinical characteristics, laboratory and radiologic findings and outcomes of the first consecutive 1096 patients who tested positive for SARS-COV-2 and were hospitalized at Jaber Al-Ahmad Al-Sabah in Kuwait.

## Materials and Methods

### Participant recruitment and study design

Jaber Al-Ahmad Al-Sabah hospital is a 1240 bed tertiary hospital based in South Surra, Kuwait. All patients admitted to Jaber Al-Ahmad Al-Sabah hospital in Kuwait, with a diagnosis of COVID-19, based on the World Health Organization (WHO) interim guidance^4^ and have been confirmed by laboratory testing using PCR testing, between February 24^th^ 2020 and the study’s cutoff date of April 20^th^ 2020 were included in the study. Patients who had equivocal testing results or were suspected cases were excluded from the study. Ethical approval for this study was obtained from the Kuwait Ministry of Health Ethical Review Board.

### Data collection

Data regarding patients’ demographics and initial clinical presentation (signs, symptoms, laboratory and radiographic findings) was collected for the study from Jaber Al-Ahmad Al-Sabah hospital’s electronic medical record system. Radiologic findings were extracted from the radiologists’ reports on the electronic medical record system. A custom data collection form was created using the SurveyCTO (Dobility, Inc) platform. To minimize data entry errors, appropriate constraints were placed on most of the data entry fields. All data then underwent a secondary quality check and was reviewed by a physician and a statistician. Any discrepancies were resolved by a third physician to ensure accuracy of the data that was entered.

### Definitions

Study participants’ signs, symptoms, vital signs, laboratory investigations and radiologic findings were all recorded on admission. ‘On admission’, ‘at the time of diagnosis’ and ‘on presentation’ are used interchangeably to describe the variables that were collected initially for patients on the first day they were admitted to hospital. All the chest x-rays that were ordered on admission for patients were reviewed by a consultant radiologist and formally reported. The normal reference ranges for vital signs are provided in (Appendix 1). Fever was defined as an oral temperature of 38.0°C and above. The disease severity of Covid-19 on admission was quantified using the quick sequential organ failure assessment score (qSOFA) score for sepsis^5^. Patients’ BMIs^6^ were categorized according the WHO criteria. The diagnostic criteria that were used for the adverse events that were collected are listed in (Appendix 2). The treatment protocols of patients with COVID-19 pneumonia at our center followed the official Kuwait Ministry of Health COVID-19 management guidelines, the latest version of which can be found in (Appendix 4). All patients diagnosed with COVID-19 stayed in the hospital until they had resolution of symptoms; defined as being afebrile for more than 72 hours and having oxygen saturations equal to or above 94%, Discharge occurred after two consecutive negative PCR tests for COVID-19, more than 24 hours apart.

### Laboratory Investigations

All diagnostic tests were performed in Jaber Al-Ahmad Al-Sabah hospital in Kuwait. COVID-19 testing was performed via real-time reverse-transcriptase-polymerase chain-reaction (RT-PCR) assay of specimens obtained via nasopharyngeal swabs.

### Statistical Analysis

Entered data were checked for accuracy, then for normality, using Kolmogorov-Smirnov & Shapiro-Wilk tests, and proved to be not normally distributed. Qualitative variables were expressed as numbers and percentages while quantitative variables were expressed as medians and interquartile ranges (IQR). Vital signs were categorized as ‘normal’ if their value was within the normal reference range, as reported in Appendix 1., and ‘abnormal’ if their value was below or above the normal range. Laboratory values were categorized as ‘below’, ‘within’ and ‘above’ the reference ranges described in Appendix 3. Clinical outcomes were subcategorized by age: below 18, between 18 and 64 and above 65 years old. For the possible confounding effects of the variables, multiple logistic regression were used for the final analysis to predict factors which may be associated with COVID-19 outcomes; namely, mortality and admission to the intensive care unit (ICU). All the explanatory variables included in the logistic models were categorized into two levels (0 for no and 1 for yes). In multivariate analysis, the associations between exposure and outcomes were expressed in terms of odds ratio (OR), together with 95% confidence intervals (95% CI). A 5% level is chosen as a level of significance in all statistical significance tests used. All statistical analysis was performed using IBM SPSS® version 22 for Windows.

### Role of Funding

A research grant was awarded by the Kuwait Foundation for the Advancement of Science to aid in data collection, and purchasing statistical software and database packages for the paper.

## Results

### Demographics and Baseline Characteristics

A total of 1096 patients were included in the study. Table 1 summarizes our study sample’s demographics, recent travel history and source of COVID-19 transmission. The median age of our sample was 41 years (inter-quartile range, 25-57 years old). Of those patients, 888 (81%) were male. Most patients were of Indian origin (48·1%), followed by Kuwaitis (27·1%) and Egyptians (6·6%). Of note, 506/527 (96·0%) of the Indian nationals were male. The mean BMI of our sample was 26·6 (SD, 8·3), after excluding children below the age of 12. 41·5% of those were classed as overweight, with a BMI between 25-29·9. Most of the patients were non-smokers 1052/1096 (96·0%). Many patients had a history of recent travel (within the past 30 days), 287/1096 (26·2%); mainly from the United Kingdom (8·7%) and Iran (7·4%). The most common mode of COVID-19 transmission in our patient population was contact with a known positive case of COVID-19 (48·3%).

**Table 1.** Demographic Information.

|  |  |
| --- | --- |
| Total No. | 1096 |
| Age (years), median (IQR) [range]<br>Distribution – no. (%)<br>0 – 14 years<br>15-49 years<br>50 – 64 years<br>≥ 65 years | 41 (25-57) [1-93]<br>33 (3.0)<br>752 (68.6)<br>231 (21.1)<br>80 (7.3) |
| Sex – no. (%)<br>Male<br>Female | 888 (81.0)<br>208 (19.0) |
| Height (cm), mean [SD]<br>Weight (kg), mean [SD]<br>BMI (kg/m<sup>2</sup>), mean [SD]<br>WHO criteria BMI cut off– no./total<br>no. above age 12 years old (%)<br>Underweight (<18.5)<br>Normal (18.5 -24.9)<br>Overweight (25 – 29.9)<br>Obese (≥ 30) | 166.0 [8.3]<br>74.9 [14.4]<br>26.6 [4.6]<br>11/674 (1.6)<br>236/674 (35.0)<br>280/674 (41.5)<br>147/674 (21.8) |
| Nationality – no. (%)<br>India<br>Kuwait<br>Egypt<br>Bangladesh<br>Iran<br>Philippines<br>European<br>Saudi Arabia<br>Other | 527 (48.1)<br>297 (27.1)<br>72 (6.6)<br>70 (6.4)<br>23 (2.1)<br>20 (1.8)<br>4 (0.4)<br>3 (0.3)<br>80 (7.3) |
| Smoker – no. (%)<br>Yes<br>No | 44 (4.0)<br>1052 (96.0) |
| Recent Travel History<br>(within past 30 days) – no. (%) |  |
| United Kingdom | 95 (8.7) |
| Iran | 81 (7.4) |
| Other (Asia) | 30 (2.7) |
| Other (Europe) | 27 (2.5) |
| France | 18 (1.6) |
| Other (Africa) | 11 (1.0) |
| United States of America | 8 (0.7) |
| Saudi Arabia | 5 (0.5) |
| Italy | 4 (0.4) |
| Spain | 4 (0.4) |
| China | 2 (0.2) |
| Other (South America) | 2 (0.2) |
| Transmission Source – no. (%) |  |
| Contact with a known positive case | 529 (48.3) |
| Unknown | 313 (28.6) |
| Travel | 238 (21.7) |
| Hospital acquired in a health care worker | 16 (1.5) |

### Clinical Characteristics on Initial Hospital Presentation

Almost half of the patients were asymptomatic at the time of diagnosis, and around 70% had no signs of infection. For the patients who presented with symptoms, the vast majority had either cough (29%), chills (26·8%) or sore throat (11·7%). Table 2 provides a summary of the symptoms present at the time of diagnosis. The vital signs taken on presentation were within normal limits for most patients, with 1039 (94·6%) having no fever initially (Table 2). Although most patients did not have any co-morbidities (69·4%), hypertension and diabetes mellitus were present in 177/1096 (16·1%) and 155/1096 (14·1%) of the patients respectively (Table 2). Of note there were 3 (0·3%) patients who were pregnant. Most patients had a qSOFA score of 0 on admission (63%) and only a small number of patients required vasopressors or intubation on admission (0·6% and 0·9% respectively) (Table 2).

**Table 2.** Clinical Characteristics on Initial Hospital Presentation.

|  |  |
| --- | --- |
| Total No. | 1096 |
| Symptoms – no. (%) |  |
| None | 543 (49.5) |
| Cough | 318 (29.0) |
| Chills | 294 (26.8) |
| Sore throat | 128 (11.7) |
| Myalgias/arthralgias | 75 (6.8) |
| Headache | 70 (6.4) |
| Fatigue | 38 (3.5) |
| Shortness of breath | 30 (2.7) |
| Nasal congestion | 27 (2.5) |
| Sputum production | 24 (2.2) |
| Nausea and/or vomiting | 18 (1.6) |
| Diarrhea | 16 (1.5) |
| Conjunctival congestion | 1 (0.1) |
| Hemoptysis | 1 (0.1) |
| Anosmia | 1 (0.1) |
| Other | 44 (4.0) |
| Duration of symptoms prior to initial presentation (days), median (IQR) [range] | 3 (0-6) [1-21] |
| Signs of infection – no. (%) |  |
| None | 761 (69.4) |
| Throat congestion | 27 (2.5) |
| Lymphadenopathy | 0 (0.0) |
| Tonsillar swelling | 0(0.0) |
| Other | 39 (3.6) |
| Comorbidities – no. (%) |  |
| None | 761 (69.4) |
| Hypertension | 177 (16.1) |
| Diabetes mellitus | 155 (14.1) |
| Dyslipidemia | 65 (5.9) |
| Asthma | 43 (3.9) |
| Coronary artery disease/ischemic heart disease | 41 (3.7) |
| Hypothyroidism | 25 (2.3) |
| Cancer | 14 (1.3) |
| Chronic renal disease | 11 (1.0) |
| Cerebrovascular disease | 7 (0.6) |
| Hepatitis | 6 (0.5) |
| Chronic obstructive pulmonary disease | 5 (0.5) |
| Recent surgery (30 days prior to initial presentation) | 4 (0.4) |
| Immunodeficiency | 1 (0.1) |
| Other | 32 (2.9) |
| Pregnancy – no. (%) | 3 (0.3) |
| Vital signs |  |
| Heart rate (beats per minute) |  |
| Mean [SD] | 86.0 [12.5] |
| Abnormal no./total no. (%) | 137/1090 (12.5) |
| Normal no./total no. (%) | 953/1090 (87.0) |
| Systolic blood pressure (mm Hg) |  |
| Mean [SD] | 127.9 [16.5] |
| Abnormal no./total no. (%) | 386/1085 (35.2) |
| Normal no./total no. (%) | 699/1085 (63.8) |
| Diastolic blood pressure (mm Hg) |  |
| Mean [SD] | 79.1 [9.8] |
| Abnormal no./total no. (%) | 402/1085 (36.7) |
| Normal no./total no. (%) | 683/1085 (62.3) |
| Respiratory rate (breaths per minute) |  |
| Mean [SD] | 20.8 [1.8] |
| Abnormal no./total no. (%) | 383/1087 (34.9) |
| Normal no./total no. (%) | 704/1087 (64.2) |
| Temperature (°C) |  |
| Mean [SD] | 36.9 [0.5] |
| Abnormal no./total no. (%) | 59/1096 (5.4) |
| ≥38°C | 57/1096 (5.2) |
| Normal no./total no. (%) | 1039/1096 (94.6) |
| Glasgow Coma Scale |  |
| Abnormal no./total no. (%) | 9/1096 (0.8) |
| Normal no./total no. (%) | 1087/1096 (99.2) |
| qSOFA score - no./total no. (%) |  |
| 0 | 691/1096 (63.0) |
| 1 | 372/1096 (33.9) |
| 2 | 33/1096 (3.0) |
| Vasopressors required on admission - no./total no. (%) |  |
| Yes | 7/1096 (0.6) |
| No | 1089/1096 (99.4) |
| Intubation required on admission - no./total no. (%) |  |
| Yes | 9/1096 (0.8) |
| No | 1087/1096 (99.2) |

### Laboratory Findings on Admission

Table 3 summarizes the laboratory findings on admission, with most patients having results within the normal laboratory reference range. Of note, 89% of the patients had a normal white blood count. Only 14·6% had a lymphocyte percentage below the reference range and 30·7% had a monocyte percentage that is above the reference range on the white blood count differential. One third of the patients were hyperglycemic on admission, and the predominant electrolyte imbalances were hyponatremia (28·7%) and hypocalcemia (20·6%). With regards to the inflammatory markers, 36·6% of patients had elevated C-reactive protein (CRP) initially and 60% had a D-dimer level lower than the normal range. More than half (53·4%) of the patients had low prothrombin time (PT) and most patients had a normal coagulation profile. In terms of liver enzymes, the aspartate transaminase (AST), was the most elevated above the normal reference range value for our sample (14·4% of patients). Most patients had albumin and total protein levels below the normal reference range (18·2% and 18·7%, respectively).

**Table 3.** Laboratory Findings on Initial Hospital Presentation.

| **Table 3.**<br>**Laboratory Findings on Initial Hospital Presentation** |  |  |  |  |
| --- | --- | --- | --- | --- |
| **Laboratory tests** | **Median (IQR)** | **Proportion of values **below** reference range no./total no. (%)** | **Proportion of values **within** reference range no./total no. (%)** | **Proportion of values **above** reference range no./total no. (%)** |
| **Complete blood count** |  |  |  |  |
| White blood count (x 10<sup>9</sup>/L) | 88 (4.6-7.2) | 88/1088 (8.0) | 975/1088 (89.0) | 25/1088 (2.3) |
| Differential |  |  |  |  |
| Neutrophils (%) | 57 (48-66) | 130/1088 (11.9) | 844/1088 (77.0) | 114/1088 (10.4) |
| Lymphocytes (%) | 31.4 (23.5-39.2) | 160/1088 (14.6) | 795/1088 (72.5) | 133/1088 (12.1) |
| Monocytes (%) | 9.1 (7.2-11.6) |  | 751/1088 (68.5) | 337/1088 (30.7) |
| Eosinophils (%) | 0.9 (0.2-2.1) | 171/1088 (15.6) | 881/1088 (80.4) | 36/1088 (3.3) |
| Hemoglobin (g/L) | 145 (133-155) | 208/1088 (19.0) | 848/1088 (77.4) | 32/1088 (2.9) |
| Red cell distribution width (RDW) (%) | 12.9 (12.3-13.6) | 95/1088 (8.7) | 1009/1088 (92.1) | 79/1088 (7.2) |
| Platelets (No.) | 229 (186-277) |  | 977/1088 (89.1) | 16/1088 (1.5) |
| **Biochemical analysis and renal function test** |  |  |  |  |
| Glucose (mmol/L) | 5.59 (4.94 – 6.8) | 26/1028 (2.4) | 636/1028 (58.0) | 366/1028 (33.4) |
| Urea (mmol/L) | 3.5 (2.8-4.3) | 169/1088 (15.4) | 868/1088 (79.2) | 51/1088 (4.7) |
| Creatinine (μmol/L) | 75 (64-86) | 501/1088 (45.7) | 551/1088 (50.3) | 36/1088 (3.3) |
| Estimated glomerular filtration rate (eGFR) (mL/min/1.73<sup>2</sup>) | 104 (92-116) | 38/1047 (3.5) | 1009/1047 (92.1) |  |
| Magnesium (mmol/L) | 0.82 (0.78-0.87) | 97/982 (8.9) | 877/982 (80.0) | 8/982 (0.7) |
| Sodium (mmol/L) | 137 (135-139) | 315/1085 (28.7) | 768/1085 (70.1) | 2/1085 (0.2) |
| Potassium (mmol/L) | 4.2 (3.9-4.4) | 57/1087 (5.2) | 1019/1087 (93.0) | 11/1087 (1.0) |
| Adjusted calcium (mmol/L) | 2.27 (2.21 – 2.35) | 202/982 (20.6) | 774/982 (78.8) | 6/982 (0.6) |
| HCO<sub>3</sub> (meq/L) | 25 (24-27) | 19/279 (6.8) | 256/279 (91.8) | 4/279 (1.4) |
| **Inflammatory markers** |  |  |  |  |
| C-reactive protein (CRP) (mg/L) | 6 (3-16) | 6/1056 (0.5) | 649/1056 (59.2) | 401/1056 (36.6) |
| Procalcitonin (ng/mL) | 0.05 (0.03 – 0.08) | 2/1030 (0.2) | 939/1030 (85.7) | 89/1056 (8.1) |
| D-Dimer (mg/mL) | 199 (147-306) | 105/206 (60.0) | 71/206 (34.4) | 30/206 (14.6) |
| **Coagulation profile** |  |  |  |  |
| Prothrombin time (PT) (seconds) | 13.1 (12.5-13.7) | 391/731 (53.4) | 327/731 (44.7) | 13/731 (1.8) |
| Activated partial thromboplastin time (APTT) (seconds) | 31.2 (29.6-32.9) | 3/723 (0.4) | 678/723 (93.8) | 42/723 (5.8) |
| International Normalized Ratio (INR) | 1.02 (0.92-0.97) | 0/732 (0.0%) | 684/732 (93.4) | 48/732 (6.6) |
| **Liver function test** |  |  |  |  |
| Alanine transaminase (ALT) (IU/L) | 26 (18-38) | 14/1078 (1.3) | 968/1078 (88.3) | 96/1078 (8.8) |
| Aspartate transaminase (AST) (IU/L) | 26 (21-34) | 2/1074 (0.2) | 914/1074 (83.4) | 158/1078 (14.4) |
| Alkaline phosphatase (ALP) (IU/L) | 64 (52-76) | 289/1082 (26.4) | 749/1082 (68.3) | 44/1082 (4.0) |
| Gamma glutamyl transpeptidase (GGT) (IU/L) | 22 (15-24) | 137/1009 (12.5) | 790/1009 (72.1) | 82/1009 (7.5) |
| Total bilirubin (μmol/L) | 12 (9-15) | 9/1081 (0.8) | 962/1081 (87.8) | 110/1081 (10.0) |
| Direct bilirubin (μmol/L) | 2.2 (1.5-3.2) | 42/1081 (3.8) | 976/1081 (89.1) | 63/1081 (5.7) |
| Albumin (g/L) | 39 (36-42) | 200/1089 (18.2) | 871/1089 (79.5) | 18/1089 (1.6) |
| Total Protein (g/L) | 69 (64-72) | 205/1011 (18.7) | 799/1011 (72.9) | 7/1011 (0.6) |

### Initial Radiographic Findings

As depicted in Table 4, only one third of patients had a chest x-ray that was reported as being ‘normal’ by the radiologist on admission (33%). The most reported findings by the radiologists were prominent broncho-vascular markings (43·3%), unilateral local patchy shadowing or opacification (16·6%) or diffuse opacification (8·2%) (Table 4). Only 71 patients had a computed tomography (CT) of the chest, of which 54 (76·1%) were reported as normal by the radiologist. The most reported finding on CT chest was ground glass opacity (15·4%)

**Table 4.** Radiographic Findings on Initial Hospital Presentation.

|  |  |
| --- | --- |
| Abnormalities on chest radiograph – no. (%) | **Total = 1096** |
| None | 362 (33) |
| Prominent broncho-vascular markings | 475 (43.3) |
| Unilateral local patchy shadowing or opacification | 182 (16.6) |
| Diffuse opacification or bilateral patchy shadowing | 90 (8.2) |
| Consolidation | 17 (1.6) |
| Ground glass opacity | 3 (0.3) |
| Pleural effusion | 2 (0.2) |
| Interstitial abnormalities | 1 (0.1) |
| Cardiomegaly | 5 (0.5) |
| Other | 17 (1.6) |
| Abnormalities on chest CT scan – no. (%) | **Total = 71** |
| None | 54 (76.1) |
| Ground glass opacity | 11 (15.4) |
| Local patchy shadowing or opacification | 2 (2.8) |
| Other | 4 (5.6) |

### Treatments and Adverse Events During Hospital Admission

Based on the previously described treatment protocols at our center, a total of 27·9% of patients did not receive any medications or treatments and were admitted to hospital for institutional quarantine purposes. Most patients (68·2%) were labeled as having received ‘other treatments’, and this mostly included supportive medications such as paracetamol and vitamin supplements (Table 5). The most common non-supportive medications that were prescribed, were antibiotics (27·9%), followed by antivirals (13·8%) and hydroxychloroquine (7·1%). For therapeutic interventions, admission to the intensive care unit (3·6%), oxygen therapy (3·2%) and mechanical ventilation (2·8%) were the most frequently administered. Most patients (90·1%) had no adverse events during hospital admission. Pneumonia was the most common complication associated with COVID-19 (7·1%), followed by acute respiratory distress syndrome (7·1%) (Table 5).

**Table 5.** Treatments, Adverse Events and Clinical Outcomes During Hospital Admission.

|  |  |
| --- | --- |
| Treatments – no. (%)<br>None<br>Antibiotics<br>Antivirals<br>Hydroxychloroquine<br>Admission to intensive care unit<br>Oxygen therapy<br>Mechanical ventilation<br>Systemic glucocorticoids<br>Antifungals<br>Extracorporeal membrane oxygenation (ECMO)<br>Continuous renal replacement therapy<br>Non-invasive ventilation<br>Other | **Total = 1096**<br>306 (27.9)<br>151 (13.8)<br>78 (7.1)<br>45 (4.1)<br>40 (3.6)<br>35 (3.2)<br>31 (2.8)<br>13 (1.2)<br>8 (0.7)<br>8 (0.7)<br>5 (0.5)<br>4 (0.4)<br>748 (68.2) |
| Adverse events – no. (%)<br>None<br>Physician-diagnosed pneumonia<br>Acute respiratory distress syndrome<br>Acute kidney injury<br>Septic shock<br>Disseminated intravascular coagulation<br>Encephalopathy/encephalitis<br>Other | **Total = 1096**<br>987 (90.1)<br>78 (7.1)<br>31 (2.8)<br>14 (1.3)<br>7 (0.6)<br>3 (0.3)<br>1 (0.1)<br>26 (2.4) |
| Clinical outcomes at study cutoff date – no. (%)<br>Discharged alive<br>Died<br>Hospitalized, in ICU<br>Hospitalized being actively treated<br>Hospitalized not receiving active treatment | **Total = 1096**<br>967 (88.2)<br>19 (1.7)<br>19 (1.7)<br>26 (2.4)<br>65 (5.9) |
| Length of hospital stay – median, (IQR) [range]<br>For all patients<br>By age<br><18 years old<br>18-65 years old<br>>65 years old | 18 (13-24) [2-64]<br>17 (13-20) [3-41]<br>17 (13-23) [2-64]<br>24 (18-31) [8-64] |

### Clinical Outcomes

At the study’s cutoff date, most of the patients in our cohort were eventually discharged from the hospital (88·2%), and only 19 (1·7%) patients succumbed to the disease. The median length of stay (LOS) for the patients was 18 days (interquartile range, 13-24 days), with patients aged above 65 years having the longest median LOS of 24 days (interquartile range, 18-31 days). A breakdown by age of the most frequent adverse events and clinical outcomes is outlined in Table 6.

On multivariate analysis, the risk factors significantly associated with an admission to the intensive care unit were found to be age above 50 years old (OR: 2·88 [95% CI, 1·05-7·95], *p*=0·041), a qSOFA score above 0 (OR: 2·80 [95% CI, 1·25-6·26], *p*=0·012), smoking (OR: 5·86 [95% CI, 1·40-24·47], *p*=0·015), elevated CRP levels (OR: 9·08 [95% CI, 1·97-41·95], *p*=0·005) and elevated procalcitonin levels (PCT) (OR: 7·00 [95% CI, 2·79-17·59], *p*=0·00003). As for mortality risk, the factors that were found to have a significant associations were asthma (OR: 4·92 [95% CI, 1·03-23·44], *p*=0·046), smoking (OR: 10·09 [95% CI, 1·22-83·40], *p*=0·032 and elevated PCT (OR: 8·24 [95% CI, 1·95-43·74], *p*=0 004). (Table 7).

**Table 6.**
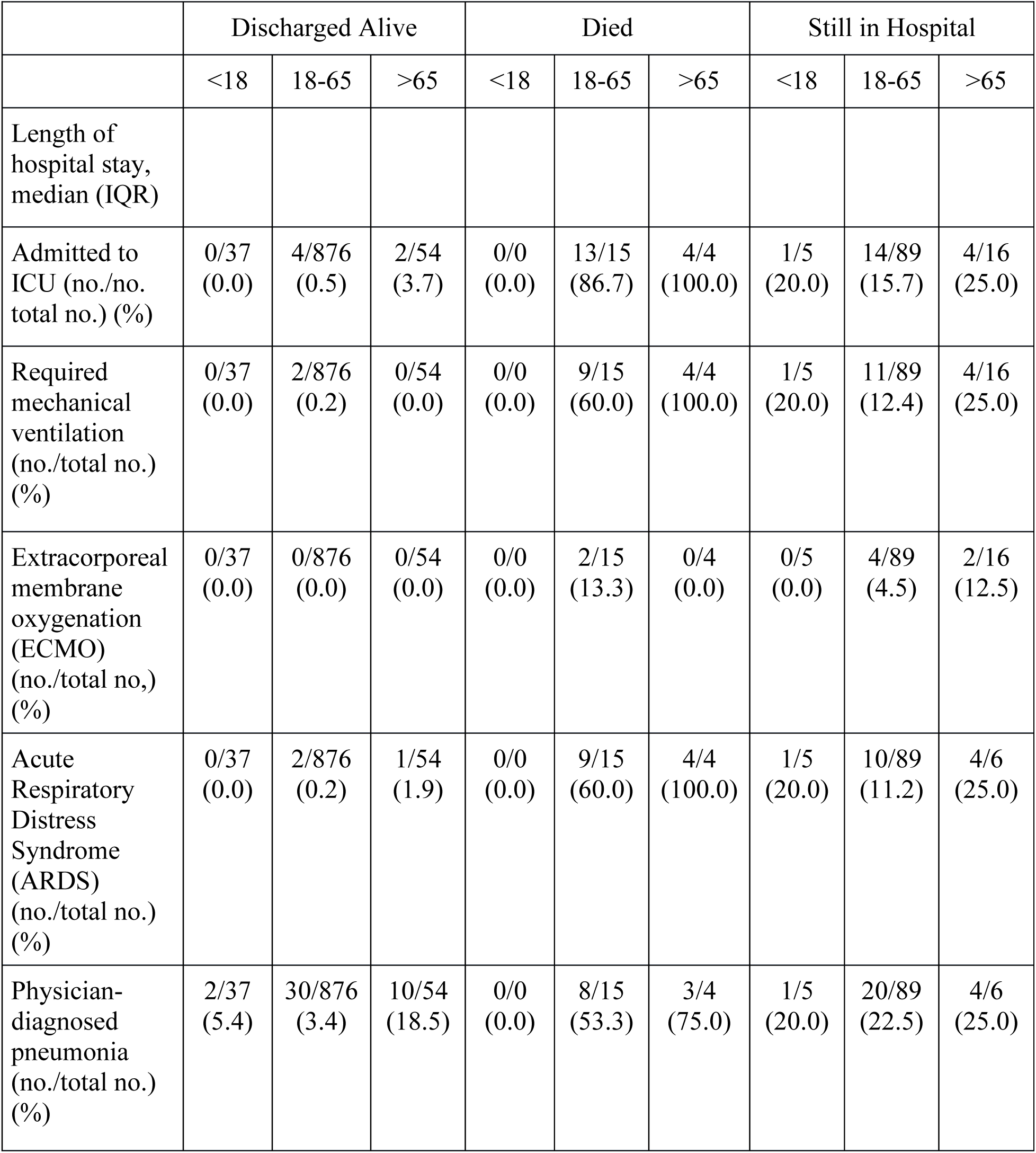
Clinical outcomes for patients at study cutoff date by age.

**Table 7.** Multivariate analysis of factors associated with mortality or admission to intensive care.

|  |  |  |
| --- | --- | --- |
|  | Multivariable Odds Ratio<br>(95% CI) | *p*-value |
| Mortality |  |  |
| Asthma | 4.92 (1.03-23.44) | 0.046 |
| Smoker | 10.09 (1.22-83.40) | 0.032 |
| Elevated procalcitonin | 8.24 (1.95-34.74) | 0.004 |
| Admission to Intensive Care |  |  |
| Age >50 years old | 2.88 (1.05-7.95) | 0.041 |
| qSOFA score > 0 | 2.798 (1.25-6.26) | 0.012 |
| Smoker | 5.86 (1.40-24.47) | 0.015 |
| Elevated CRP | 9.08 (1.97-41.95) | 0.005 |
| Elevated procalcitonin | 7.00 (2.79-17.59) | 0.00003 |

## Discussion

To our knowledge, this is one of the first large retrospective cohort studies to summarize demographic, clinical characteristics and outcomes of consecutive COVID-19 patients in a single country. Our study sample had several distinguishing features, such as; a large proportion of asymptomatic hospitalized COVID-19 patients (49·5%), the majority of patients had definitive outcomes at the study’s cutoff date (90·0%) and all the patients in our sample were admitted at a single center for treatment and/or quarantine purposes where they all received standardized investigations and treatment protocols, irrespective of disease severity (Appendix 4.). In this study sample, we found an association between several risk factors and admission to the intensive care unit: namely, age above 50 years old, smoking, elevated qSOFA score, elevated C-reactive protein and procalcitonin levels. Also, the following risk factors were identified as having a correlation with mortality in our sample: asthma, smoking and elevated procalcitonin levels.

The median age for our sample was lower (41 years old), compared to the two other large retrospective cohort studies from China^1^ (47 years old) and New York City^2^ (63 years old). This is likely a result of our sample encompassing a large cohort of patients who were asymptomatic and were only identified as being COVID-19 positive due to the mass screening efforts of the Kuwaiti government for incoming travelers. This may also account for the large proportion of patients with a history of recent travel in our cohort (21·7%). Only 19% of our study’s patients were female, which is lower but in keeping with findings by Guan et al.^1^ and Richardson et al.^2^, who also reported a lower admission rate for women compared to men (41·9% and 39·7%, respectively). A contributing factor to the lower proportion of women in our study sample, may be the high rates of COVID-19 detected in manual laborer of Indian ethnicity, who tend to be both male and younger. An outbreak in two main epicenters, *Al-Jileeb* and *Mahboula* areas in Kuwait, both of which house high concentrations of Indian male manual laborers may also account for the large proportion of young, male, Indian patients in our study sample (48·1%)^7^. Also, South Asian ethnicity and lower socio-economic state have been hypothesized to be associated with higher rates of COVID-19 and poorer outcomes, based on epidemiological observations^8^. The mean BMI for our sample was 26·6 kg/m^2^, with 41·5% of patients in the overweight category, which is reflective of the normal weight demographics in Kuwait^9^.

As reported by other studies^10^, hypertension (16·1%) and diabetes mellitus (14·1%) were the most common co-morbidities in our cohort. For symptomatic patients, the most common symptom was cough (49·5%), which is in keeping with several other retrospective cohort studies^1–3,11,12^. Interestingly, almost half of our sample were asymptomatic on initial presentation (49·5%) and had no signs of infection on clinical examination (69·4%). This may explain why only 5·4% of our patient population had a temperature of 38°C or above, which is in stark contrast to the other retrospective cohort studies in COVID-19 patients which reported much higher rates (Richardson et al, 30·7%, Guan et al., 59·2%, Wu et al. 38·3% above 39°C, Zhou et al. 94% above 37·3°C), as almost all their patients were symptomatic. As a result, we believe our study sample provides a more accurate portrayal of the clinical manifestations of COVID-19 in a general population. Our findings also raise the question of whether fever is an effective screening tool for COVID-19.

Most of the laboratory findings were within the normal reference range in our study, probably secondary to the mild disease present in most of the cases. Interestingly, a large proportion of patients had sodium and calcium levels below the reference range on admission (28·7% and 20·6%, respectively). Hong et al.^13^ recently reported similar findings, where 50% of COVID-19 patients they examined had hyponatremia and hypokalemia and they suggested there was a correlation with the degree of renal injury in those patients. Similarly, Sun et al.^14^ found high rates of hypocalcemia (74·7%) in their COVID-19 patient population, on admission. Elevated C-reactive protein levels were present in a large percentage of patients (36·6%), as noted by other studies^15–17^. Further studies are needed to determine whether CRP can be used a surrogate for monitoring disease severity and resolution. Additionally, prothrombin levels below the reference range were present in over half our study population (53·4%). The implications of this findings are unclear but we hypothesize it may be related the procoagulant state that is thought to occur in COVID-19 patients^18^. As almost all our patients had a chest x-ray on admission, we determined that 76·3% patients either had a normal chest-ray or benign findings such as prominent broncho-vascular markings, as reported by a radiologist. Conversely, Wong et a. reported that in a cohort of 64 patients they studied, only 31% of COVID-19 had normal initial chest x-ray^19^. Of our patients with abnormal chest x-ray findings, 16·6% had unilateral local patchy shadowing or opacification, as the most common pathological finding.

Most of our patient cohort did not experience any adverse events (90·1%). As other studies have shown, pneumonia and ARDS were the most common complication of COVID-19^11^. As a result, the most prescribed medications were antibiotics (13·8%). Of note, 27·9% of patients received no medications and 68·2% received ‘other’ treatments. ‘Other treatments’ consisted of supportive medications such as paracetamol and ibuprofen and vitamin supplements, such as vitamin C and D, as well as prophylactic anticoagulation, as outlined in Kuwait’s Ministry of Health COVID-19 protocols (Appendix 4).

The median length of hospital stay was 18 days (IQR 13-24) for our study sample. This was longer that the median length of stay observed in similar studies^1,2^. In addition, the length of stay was longer in older patients, above 65 years of age in our sample (24 days, IQR 18-31). This may reflect the stringent discharge criteria at our center (2 consecutive nasopharyngeal swabs, 24 hours apart after symptom resolution). Despite the prolonged median length of hospital stay for most of our patients, 88·2% were discharged alive at the study cutoff date, 90·1% of patients had no reported adverse events during their admission, 3·6% required an ICU admission and only 1·7% died. This may reflect the young age of our study population and the fact that our cohort included a large portion of asymptomatic patients. Our mortality rates and admission rates are similar to Guan et al.^1^ (mortality, 1·4% and ICU admission, 5%), but much lower than the other large retrospective cohort studies (Wu et al., 21·9% mortality, 26·4% ICU admission^11^, Zhou et al., 28·3% mortality, 26% ICU admission^3^ and Richardson et al.^2^). This may reflect that the study by Guan et al., was one of the earliest COVID-19 retrospective cohort studies so the included patients may have had milder symptoms, compared to studies that were published later, when health resources became more limited.

Multivariate analysis, demonstrated an association between mortality and being a smoker, having asthma and elevated procalcitonin levels. There was also an association between ICU admission and age above 50 years old, a qSOFA above 0, smoking, elevated CRP levels and elevated procalcitonin levels. Interestingly, being a smoker and elevated procalcitonin levels were the only factors found to be correlated with both mortality and ICU admission. Both factors have also been associated with unfavorable outcomes by other recent studies^20,21^. Similarly, several studies have found a correlation between older age^2,11,22^ and elevated CRP levels^15–17^ and poor outcomes. High qSOFA and SOFA score was found to be correlated with death by Zhou et al^3^. Our findings did not find an association between qSOFA score and death, but we did find a correlation with ICU admission. This may be due to the small number of deaths in our sample. Although an association between asthma and poor outcomes has been suggested by some authors^23,24^, we did not find any other studies that have reported this.

### Limitation

Most of the limitations in our study are due to its retrospective nature, such as potential loss of data due to omissions. In addition, for asymptomatic patients, we did not follow their hospital course and they may have developed symptoms later during their admission. However, based on unpublished reports, we have observed that most patients who were asymptomatic on admission, remained asymptomatic^7^. Also, as the number of COVID-19 cases escalated, the guidelines for its treatments and discharge/diagnostic criteria slightly evolved over time, which may have implications on our results. At the end of the study, some patients remained hospitalized (10%) and their clinical course is still unclear.

### Conclusions

This study is, to our knowledge, the first comprehensive study to provide data on the initial 1097 consecutive COVID-19 cases of an entire country, all admitted to a single center, undergoing the same investigations and treatment protocols. We believe it gives a different perspective on the nature and clinical course of this novel disease, compared to other large retrospective cohort studies, as it includes patients with a wide spectrum of disease severity. Future studies, directed at further characterizing risk factors for disease severity and outcomes, particularly predictive scoring systems, are needed.

## Data Availability

All data referred to in the manuscript has been made available

## Acknowledgments

Kuwait’s National Center for Health Information for assistance with statistical analysis for this study.

## Appendix 1

<u>Normal range for vital signs:</u>

|  |  |
| --- | --- |
| Heart rate (beats per minute) | 60-100 |
| Systolic blood pressure (mm Hg) | 100-130 |
| Diastolic blood pressure (mm Hg) | 60-80 |
| Respiratory rate (breaths per minute) | 12-20 |
| Temperature | 36-37.5 |
| Glasgow coma scale | 3-15 |

## Appendix 2

Appendix 2.
| Adverse Event | Diagnostic Criteria |
| --- | --- |
| Septic shock | The third international consensus definitions for sepsis and septic shock (sepsis-3)<sup>25</sup> |
| Acute respiratory distress syndrome | WHO interim guidance: Clinical management of severe acute respiratory infection when COVID-19 is suspected<sup>4</sup> |
| Acute kidney injury | Kidney Disease Improving Global Outcomes (KDIGO) clinical practice guidelines for Acute Kidney Injury<sup>26</sup> |
| Disseminated intravascular coagulopathy | International Society on Thrombosis and Hemostasis (ISTH) criteria for Disseminated Intravascular Thrombosis (DIC)<sup>27</sup> |
| Physician-diagnosed pneumonia | American Thoracic Society and Infectious Diseases Society of America: Clinical Practice Guidelines<sup>28</sup> |
| Myocarditis/cardiomyopathy | World Heart Federation Consensus Conferences Definition of Inflammatory Cardiomyopathy<sup>29</sup> |
| Encephalitis/encephalopathy | Consensus statement of the international encephalitis consortium<sup>30</sup> |
| Rhabdomyolysis | Diagnosis is made in patients with either dark urine or an acute neuromuscular illness with an elevation of serum creatine kinase x 5 upper limit<sup>31</sup> |

## Appendix 3

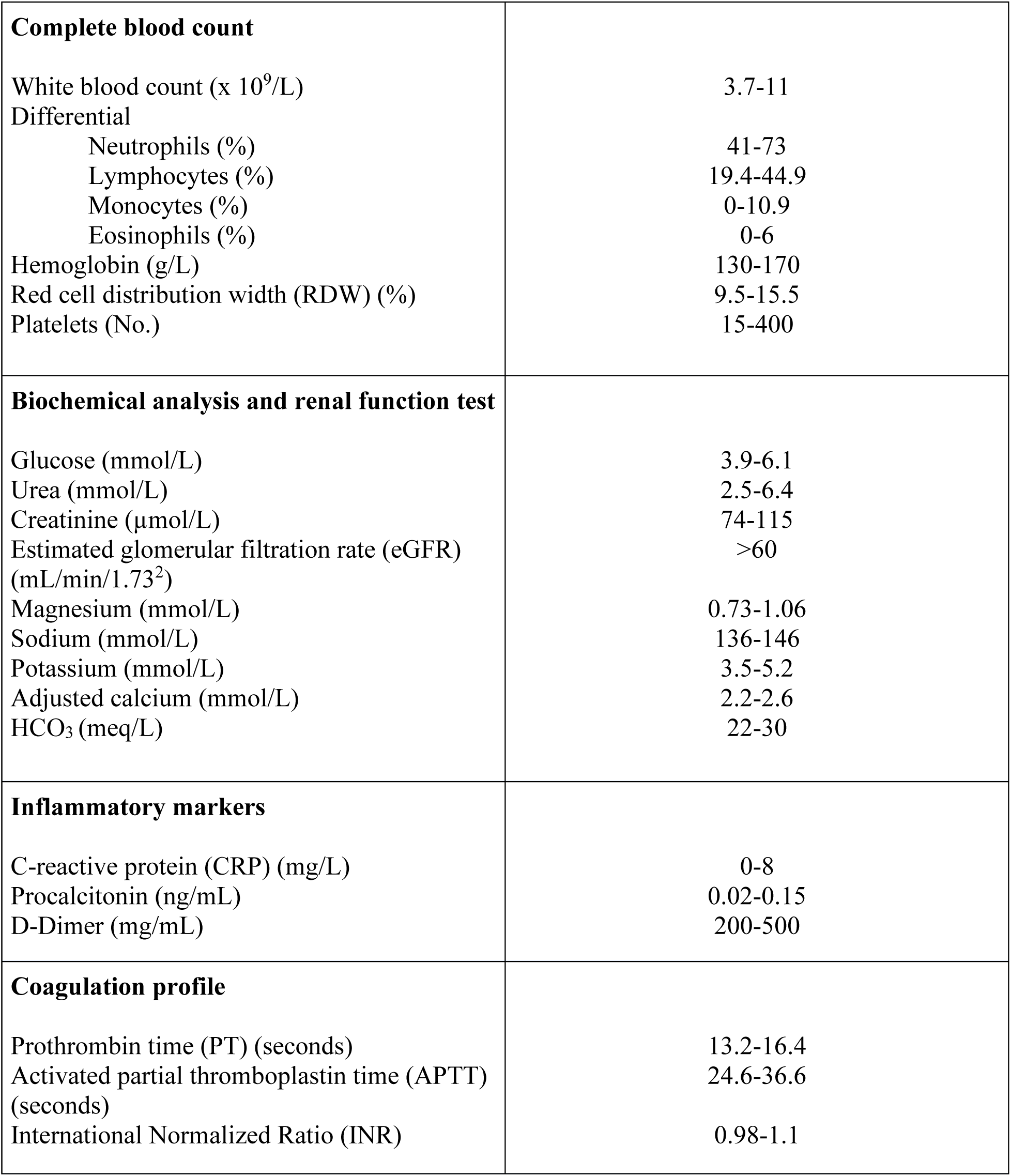
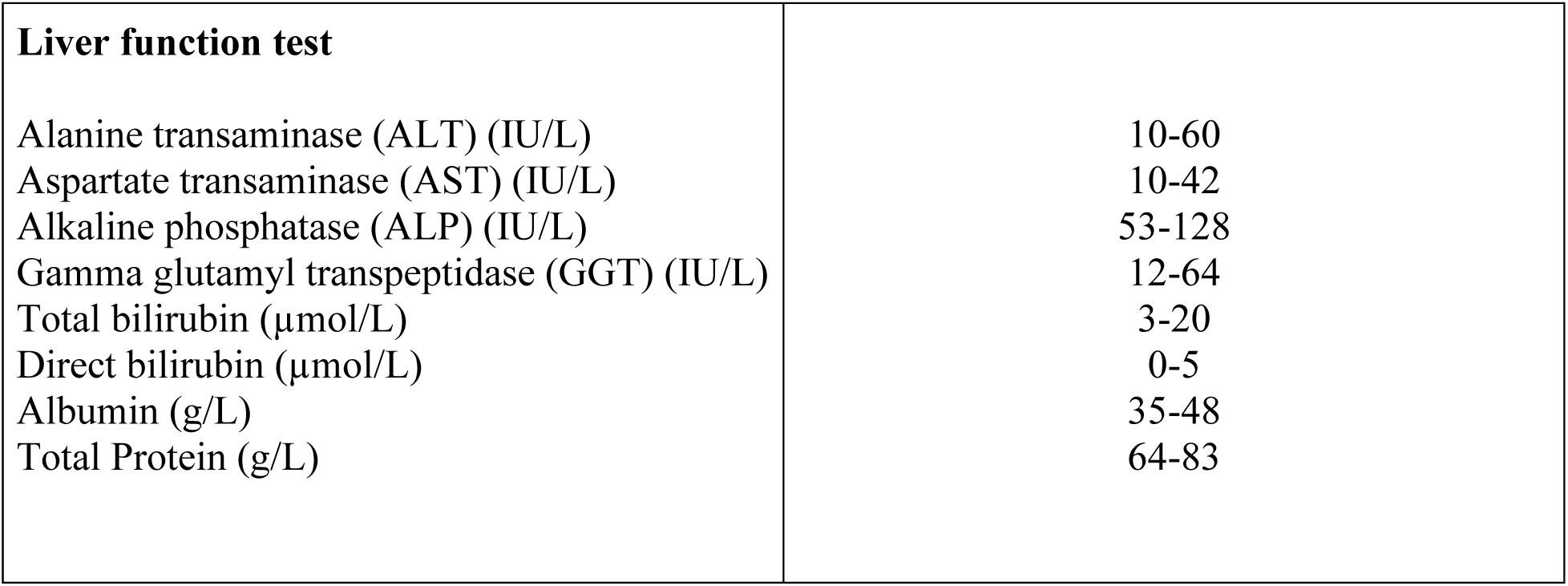

## Appendix 4

Management of Hospitalized Adult Patients with COVID-19 Pneumonia

Interim guidance: 25^th^ April 2020 (version 6)

> *“It can be ethically appropriate to offer individual patients experimental interventions on an emergency basis outside clinical trials, provided that no proven effective treatment exists; it is not possible to initiate clinical studies immediately; the patient or his or her legal representative has given informed consent; and the emergency use of the intervention is monitored, and the results are documented and shared in a timely manner with the wider medical and scientific community”*.
>
> *WHO, Off-label use of medicines for COVID-19, Scientific Brief, 31 March 2020^1^*

### 1. Definitions

Clinical syndromes associated with 2019-nCoV infection
- <u>Uncomplicated illness</u>. Patients with uncomplicated upper respiratory tract viral infection (refer to symptoms list below). The elderly and immunosuppressed may present with atypical symptoms. These patients do not have any signs of dehydration, sepsis.
- <u>Mild pneumonia</u>. Patient with pneumonia (Infiltrate on imaging + clinical symptoms consistent with pneumonia) and no signs of severe pneumonia (PSI > 130 or SIRS ≥ 2).
- <u>Severe pneumonia</u>. Fever or suspected respiratory infection, plus one of respiratory rate >30 breaths/min, severe respiratory distress, or SpO2 <90% on room air.
- <u>Sepsis</u>. Life-threatening organ dysfunction caused by a dysregulated host response to suspected or proven infection, with organ dysfunction. Signs of organ dysfunction including altered mental status, difficult or fast breathing, low oxygen saturation, reduced urine output, fast heart rate, weak pulse, cold extremities or low blood pressure, skin mottling, or laboratory evidence of coagulopathy, thrombocytopenia, acidosis, high lactate or hyperbilirubinemia.
- <u>Septic shock</u>. Persistent hypotension despite volume resuscitation, requiring vasopressors to maintain MAP ≥65 mmHg and serum lactate level >2 mmol/L.

Stratification Risk Factors by Age, Gender and Comorbidities^3–6^
- Age ≥ 60 years and older
- Male gender
- Those with comorbidities (10.5% for cardiovascular disease, 7.3% for diabetes, 6.3% for chronic respiratory disease, 6% for hypertension, and 5.6% for cancer) Plus smokers.
- Immunocompromised hosts (T-cell depleted medications; Corticosteroid equivalent to prednisone >15 mg per day for more than 2 months).

Stratification by Clinical Symptoms^7–9^
- Fever
- Dyspnea
- Fatigue
- Sore throat
- Approximately 90% of patients present with more than one symptom, and 15% of patients present with fever, cough, and dyspnea^10^
- Patients may present with nausea or diarrhea 1 to 2 days prior to onset of fever and breathing difficulties^11–12^.
- Sudden loss of taste and smell may present as an early symptom^13^.
- Cough (usually dry)
- Myalgia
- Anorexia

Stratification based on Lab/Imaging Abnormalities^14–15^
- lymphocytopenia and leukopenia
- Thrombocytopenia
- Elevated CRP
- Troponin Leak
- Acute Kidney Injury (rising creatinine)
- Elevated D-Dimer
- Elevated Ferritin
- SOFA
- Infiltrate on CXR

### 2. Initial investigations:^4,15^

**Asymptomatic/mildly symptomatic low risk patients:** perform investigations if indicated.

**Asymptomatic/mildly symptomatic high risk patients** will only need CBC, Metabolic panel, CRP, Baseline CXR, baseline ECG and G6PD level.

Order the following investigations in patients with **pneumonia/ICU**:

|  |  |  |
| --- | --- | --- |
| Pulse oximetry | ABG (if indicated) | CBC with differential |
| Comprehensive metabolic panel | Coagulation screen + D-Dimer<sup>20</sup> | Inflammatory markers (procalcitonin and CRP) |
| Serum troponin<sup>16-18*</sup> (repeat q2-3d if elevated) | Serum Vitamin D | LDH |
| Creatinine Kinase | MRSA nasal screening | Blood and Sputum Culture |
| Respiratory viral panel | Pneumonia PCR panel | G6PD |
| Ferritin<sup>19</sup> | ECG (Establish QTc baseline) | CXR |
| Triglycerides | IL-6<sup>22</sup> | Non-contrast CT is of limited <u>utility in definitively diagnosing COVID-19</u> and should only be considered if it's likely to change management in consultation with pulmonary medicine or PUI status<sup>23-24</sup> |

- **<u>If inflammatory markers (e.g. CRP) are used to monitor disease activity, it should not be repeated sooner than 3 days. Daily repeat of inflammatory markers rarely affects patient management.</u>**
- Elevated troponin (> 2 times upper limit of normal) without hemodynamic compromise, can repeat troponin in 24 hours. Up-trending troponin with hemodynamic compromise or other concerning cardiovascular symptoms signs should prompt consideration of obtaining an urgent cardiology consultation.

### 3. Management

Management principles of Uncomplicated Illness + all other patients
- Adhere to infection prevention Measures.^25^
- Vitamin C 1000 mg Tablet Daily.^26–27^
- Zinc Acétate Lozenges one every 6-8 hours.^28^
- In patients with serum Vitamin D deficiency, supplement with Vitamin D 10,000 IU daily.
- If the patient meets criteria for high risk (see appendix), consult InfectiousDiseases.
- All patients should receive standard prophylactic anticoagulation with LMWH in the absence of any contraindications^29^ (Contraindications include active bleeding or platelet count less than 25,000; monitoring advised in severe renal impairment; abnormal PT or APTT is not a contraindication. If LMWH contraindicated due to renal failure (Creatine Clearance < 30mL/min), Unfractionated Heparin 5000 units SC q12 can be used as an alternative. <u>fondaparinux</u> is preferred in those with heparin-induced thrombocytopenia.

General Management of COVID-19 Pneumonia
- In All patients requiring assistant ventilation, placement in airborne isolation is preferred.
- All patients admitted to ICU, placement in airborne isolation is preferred.
- Airborne precaution is required for all patients undergoing aerosol generating medical procedures, including nebulizer treatment. In one study, COVID19 viral particles remained viable in aerosols for 3 hours following nebulization.^30^
- If inhaler medications are needed, <u>use MDI + spacer device</u>.

#### Antimicrobial Management of COVID-19 Pneumonia

In patients < 50 yo with no comorbidities and mild pneumonia -
1. <u>ROUTINE USE OF EMPIRIC ANTIBIOTICS IS STRONGLY NOT RECOMMENDED</u>. Data has shown secondary bacterial infections are late manifestation of the disease process^31^. Further in one study, most Co-infection Between SARS-CoV-2 and Other Respiratory Pathogens are viral^32^.
2. If high suspicion for secondary bacterial infection (i.e. PCT ≥ 0.5), start second or third generation Cephalosporin or Amoxicillin-Clavulanic Acid to treat secondary bacterial infection plus Doxycycline in nonpregnant patient.
3. In PCN-allergic patients, start Moxifloxacin. Treat for 5 days only.
4. If the nasal MRSA screen is positive, consider MRSA coverage for 5 days only-Preferred agent is Teicoplanin^33^. ^34–42^
5. Consider Hydroxychloroquine^*^
6. Consult ID to consider treatment with Lopinavir-ritonavir^43^. Imperative to start treatment early. In one study late initiation of Lopinavir/ritonavir did not improve mortality rate^44^ (LPV/r is considered a second line agent)^*^.
7. Obtain CRP baseline and serially follow.
8. Obtain ECG daily, if using QTc prolonging agent.
9. **Review above antibiotic regimen in 72 hours. If started, usual course is 5 days. May discontinue if concern for bacterial pneumonia low (confirmation of COVID-19, classic presentation, PCT <0.2)**.

**\*We adopt both the Infectious Diseases Society of America (IDSA)^35^ and the American Thoracic Society (ATS)^36^ regarding COVID19 treatment chloroquine/hydroxychloroquine (HCQ), hydroxychloroquine with azithromycin, lopinavir/ritonavir, tocilizumab, corticosteroids for acute respiratory distress syndrome (ARDS). To date, there is no sufficient evidence to recommend any specific treatment. We suggest treatment on case-by-case basis using best clinical judgement. It is also important to note the French Trial *(Gautret P et al. PMID 32205204)^39^* was recently retracted by the publisher^40^**.

In patients <50 yo with comorbidities OR > 50 yo regardless of associated comorbidities, and mild pneumonia
1. **<u>ROUTINE USE OF EMPIRIC ANTIBIOTICS IS STRONGLY NOT RECOMMENDED</u>**. Data has shown secondary bacterial infections are late manifestation of the disease process^31^. Further in one study, most Co-infection Between SARS-CoV-2 and Other Respiratory Pathogens are viral^32^.
2. If high suspicion for secondary bacterial infection (ie PCT ≥ 0.5), start second or third generation Cephalosporin or Amoxicillin-Clavulanic Acid to treat secondary bacterial infection plus Doxycycline in nonpregnant patient.
3. In PCN-allergic patients, start Moxifloxacin. Treat for 5-7 days only.
4. If the nasal MRSA screen is positive, consider MRSA coverage for 5 days only-Preferred agent is Teicoplanin.
5. Consider Hydroxychloroquine^*^.
6. Consult ID to consider treatment with Lopinavir-ritonavir. Imperative to start treatment early. In one study late initiation of Lopinavir/ritonavir did not improve mortality rate (LPV/r is considered a second line agent) ^*^.
7. Obtain ECG daily, if using QTc prolonging agent.
8. **Review above antibiotic regimen in 72 hours. If started, usual course is 5 days. May discontinue if concern for bacterial pneumonia low (confirmation of COVID-19, classic presentation, PCT <0.2)**.

In patients requiring ICU admission
1. Start Piperacillin-Tazobactam plus Doxycycline in nonpregnant patient.
2. In PCN allergic patients – consult ID senior level physician.
3. If the nasal MRSA screen is positive, consider MRSA coverage for 7 days only-Preferred agent is Teicoplanin.
4. Start Hydroxychloroquine*.
5. Consult ID to consider treatment with Lopinavir-ritonavir. Imperative to start treatment early. In one study late initiation of Lopinavir/ritonavir did not improve mortality rate (LPV/r is considered a second line agent)^*^.
6. Consider initiation of Tocilizumab in consultation with critical care and ID (please refer to Tocilizumab treatment protocol)
7. **Convalescent plasma products is available for administration in Kuwait. (please refer to convalescent plasma treatment protocol)**.
8. Obtain baseline CRP, Ferritin, DIC panel, D-Dimer and serially follow.
9. Consult clinical virology to check cycle threshold (ct) value at the initiation of treatment and at 72h.
10. Unless placed on Telemetry, obtain ECG daily, if using QTc prolonging agent.
11. Review above antibiotic regimen in 72 hours.

### 4. Discharge Criteria for Hospitalized Patients with COVID19 ^22,23^

<u>-For asymptomatic/mildly symptomatic please follow MOH discharge guidance.</u>

-The following criteria should be used for patients with moderate-severe disease
1. Days since testing positive ≥ 14 days, **and**
2. Resolution of respiratory symptoms ≥ 7 days (not requiring supplemental oxygen; or pulmonary status at baseline), **and**
3. Afebrile ≥ 72 hours without anti-pyretics, **and**
4. Improving/Regression of abnormalities on imaging, **and**
5. A Negative real time PCR result collected from nasopharyngeal swabs.Testing suggested to be done on Day 12 and 13 of admission to complete the 14 days of hospitalization.
6. Discharged patients to be home quarantined imperatively for 14 days (wear a mask at home; maintain social distancing with family members, particularly elderly; resides in a single room with restroom and with good ventilation, and avoid sharing same utensils and cups).
7. Agree to maintain health monitoring with follow up visit in COVID19 Clinic (Jaber Hospital) in 2 weeks.
8. For more information, please follow public health guidance for home quarantine.

### 5. Drug monitoring

Monitoring for Hydroxychloroquine (HCQ) and renal dose adjustment
- The estimated half-life is 40 days
- Monitor for hemolytic anemia with CBC every 2 days. Post-marketing studies suggest the risk of hemolysis is very low. It is reasonable to start hydroxychloroquine in most patients while awaiting G6PD testing.
- Recommend avoid taking hydroxychloroquine with antacids. Separate administration by at least 4 hours.
- Hydroxychloroquine can be crushed.
- Probably safe in pregnancy, but benefits should be balanced with possible risks (in consultation with ID and OB/GYN).
- Most toxicities are associated with long-term use; Dizziness, headache, loss of appetite, nausea, vomiting; LFT abnormalities; Retinopathy with prolonged use (>5 years), not in the acute setting.
- CrCl <10 and hemodialysis: reduce dose to 400mg po x 1 day then 200mg po OD.
- Risk of QT prolongation: should be used with caution if other QT prolonging agent such as azithromycin or fluoroquinolones or if electrolytic imbalances (Keep potassium > 4.0 mg/dL and Magnesium > 0.82 mmol/L).
- We do not recommend co-administration with azithromycin.
- <u>Cardiac monitoring guidance</u> (***see appendix 4***)

Monitoring Parameters for Lopinavir-ritonavir (Kaletra)
- Alternative therapy if hydroxychloroquine is unavailable or if the patient has contraindications or adverse effects.
- Adverse events: Hepatotoxicity, pancreatitis, diabetes, QT prolongation, lipid elevations, and fat redistribution
- Major substrate and inhibitor of Cytochrome P450, and can cause severe drug-drug interactions. Thorough evaluation of a patient’s mediation profile and clinical pharmacy consultation should be initiated before starting therapy.
- Pregnancy: Lopinavir-ritonavir is safe to use during pregnancy.

##### Not Recommended (alphabetical order): Risk/benefit ratio does not favor use

###### Angiotensin/RAS Blocking Agents (ACEi/ARBs)^45–47^

Do not discontinue these therapies for COVID-19 disease. Multiple professional societies in cardiology and have reviewed the current data and conclude that the evidence suggesting discontinuation of ACEi/ARB therapy to decrease risk for more severe COVID-19 is not well supported at this time.

###### Azithromycin^42^

No activity for SARS-CoV-2. Single study of combination therapy with hydroxychloroquine does not convincingly suggest added benefit to azithromycin combination therapy, given the study was recently retracted and concern for antibiotic overuse.

###### Ibuprofen/NSAIDs^48^

Do not discontinue these therapies for COVID-19 disease. Paracetamol is the preferred fever reducer for use in COVID-19. Although there has been theoretical concern raised for these agents worsening outcomes, no data currently exist to support this. FDA is not aware of scientific evidence connecting the use of NSAIDs, like ibuprofen, with worsening COVID-19 symptoms. The agency is investigating this issue further and will communicate publicly when more information is available.

###### Corticosteroids^49–52^

There is significant interest and controversy surrounding the role of corticosteroids for the management of severe pneumonia due to coronaviruses. <u>The potential benefit of these agents to blunt the inflammatory cascade seen in severe disease needs to be carefully weighed against the concerns for secondary infections</u>, adverse events, and other complications of corticosteroid therapy. The data for corticosteroids are inconsistent, confusing, and inconclusive. While target patients where corticosteroids will improve outcomes may exist (e.g., those with cytokine-related lung injury who may develop rapidly progressive pneumonia), <u>that population remains ill-defined</u>. Clinicians need to carefully weigh the risks and benefits of corticosteroids on the individual patient level. This need for a risk benefit assessment in individual patients and careful consideration of dose is exemplified in the COVID-19 Diagnosis and Treatment Guide from the National Health Commission of the People’s Republic of China where the authors state ***"Based on respiratory distress and chest imaging, may consider glucocorticoid that is equivalent to methylprednisolone 1-2 mg/kg/day for 3-5 days or less. Note that large-dose glucocorticoid suppresses immune system and could delay clearance of SARS-CoV-2.”*** <u>A recent consensus statement from the Chinese Thoracic Society recommends a lower dose, ≤0.5-1 mg/kg/day methylprednisolone for ≤ 7 days in select patients,</u> after careful consideration of risks and benefits.

###### Tocilizumab^53–55^

IL-6 inhibitor; IV formulation. Preliminary clinical experience from China in COVID-19 reported benefit, however neutropenia can be long-lasting so risk of secondary infection is possible and unquantified.

### 6. List of Appendices

- **Appendix 1:** Flow sheet for stratifying risk factors to initiate infectious diseases referral
- **Appendix 2:** High risk features
- **Appendix 3:** Brief Overview of Agents
- **Appendix 4:** Cardiac Monitoring for patients on Hydroxychloroquine.

### Members of the COVID-19 infectious disease task force

Dr. Wasl Al-Adsani, Dr. Khalid Alothman, Dr. Mohammad Alghounaim, Dr. Almonther Alhasawi, Dr. Kelly Schrapp, Dr. Osama Albaqsami, Dr. Mohammed Saraya, and Dr. Ghanem Alhojailan

**Appendix 1:**
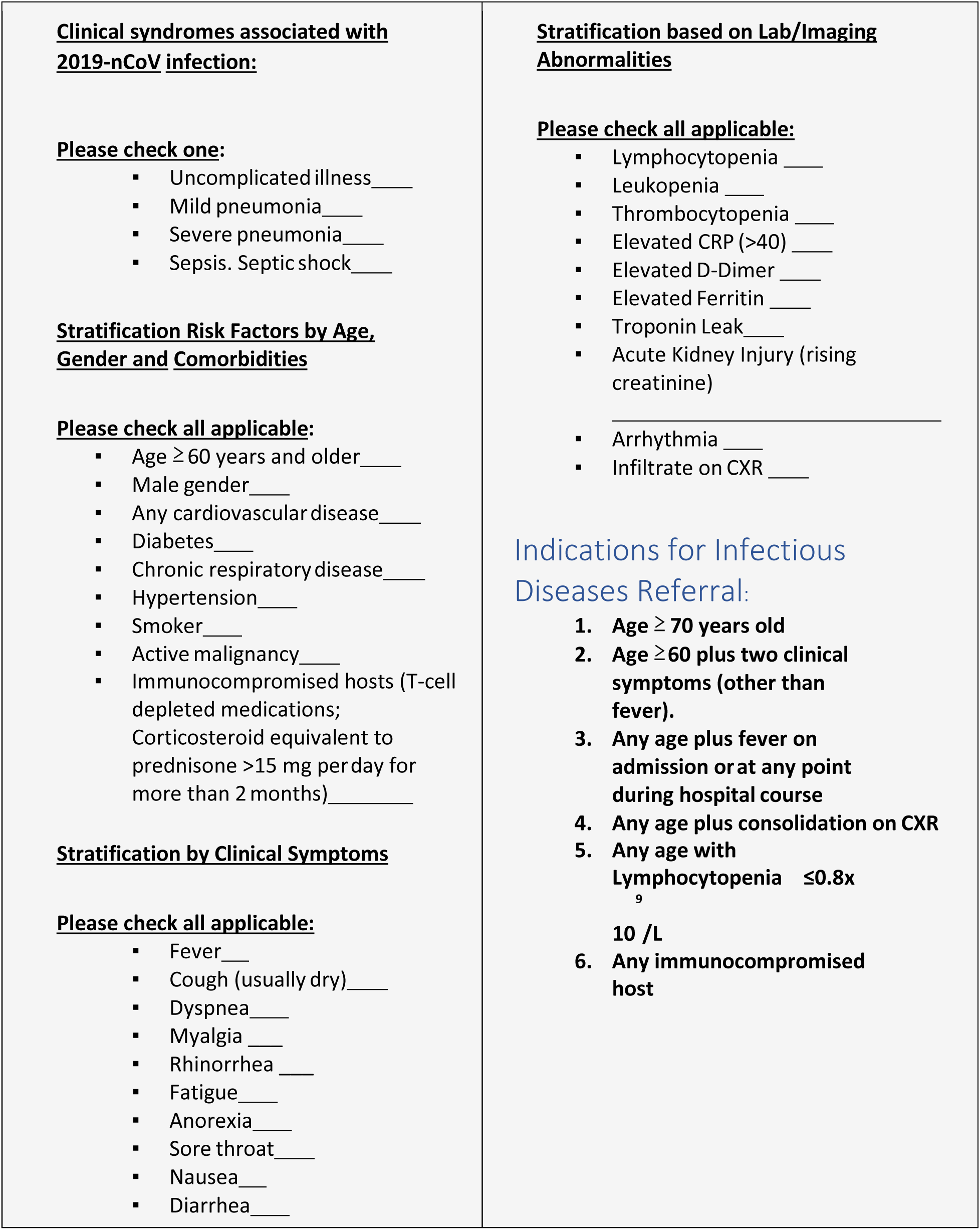
Flow sheet for stratifying risk factors to initiate infectious diseases referral^56–60^

**Appendix 2:** High Risk Features^60^:

| Epidemiological Category | Vital Signs | Labs |
| --- | --- | --- |
| Age > 50 | Respiratory rate > 24 breaths/min | D-dimer > 1000 ng/ml |
| Pre-existing pulmonary disease | Heart rate > 125 beats/min | CPK > twice upper limit of normal |
| Chronic Kidney disease | SpO2 <90% on ambient air | CRP > 60 |
| Diabetes with Alc > 7.6% |  | LDH > 345 U/L |
| History of hypertension |  | Elevated troponin |
| History of Cardiovascular disease |  | Admission absolute lymphocyte count < 1.0 |
| Use of biologics |  | Ferritin > 300 ug/L |
| History of transplant or other immunosuppression |  |  |
| All patients with HIV (regardless of CD4 count) |  |  |

**Appendix 3:** Brief Overview of Agents:

| Agent | Classification | Target/Mechanism | Dosing | Key Toxicities |
| --- | --- | --- | --- | --- |
| Hydroxychloroquine (Plaquenil) | Off-label | Multiple actions: prevents binding to ACE2; prevents transport in endosome; and possibly others | 400 mg PO BID x 2 doses (loading) then 200 mg BID for 5 days. | QTc prolongation; |
| Lopinavir/ritonavir (LPV/r or Kaletra) | Off-label | Protease inhibitor | 400/100 mg BID for 6-10 days | QTc prolongation; ALT elevation; multiple drug-drug interactions |
| Teicoplanin (Targocid) | S. aureus infections | Cell wall antibacterial agent | 6 mg/kg IV q12 x 3 loading doses; then 6 mg/kg IV q daily | Thrombocytopenia; red man syndrome; fever |
| Piperacillin-tazobactam | Broad spectrum antibacterial | Cell wall antibacterial agent | 4.5 mg IV q8 | Possible increase AKI, allergy, thrombocytopenia |
| Levofloxacin | Community acquired pneumonia | Fluoroquinolone, DNA replication inhibitor | 500 mg IV/PO daily | Neurotoxicity, Qt prolongation, tendinopathy |
| Tocilizumab | Off-label | IL-6 blocker | Single dose 4 – 8 mg/kg (usual dose 400 mg; maximum 800 mg) by IV infusion; repeated within 12 hours for maximum of 2 doses | Hepatitis, leukopenia, HSV reactivation, TB reactivation |

**Liverpool COVID-19 Drug Interactions: http://www.covid19-druginteractions.org/**

**Appendix 4:**
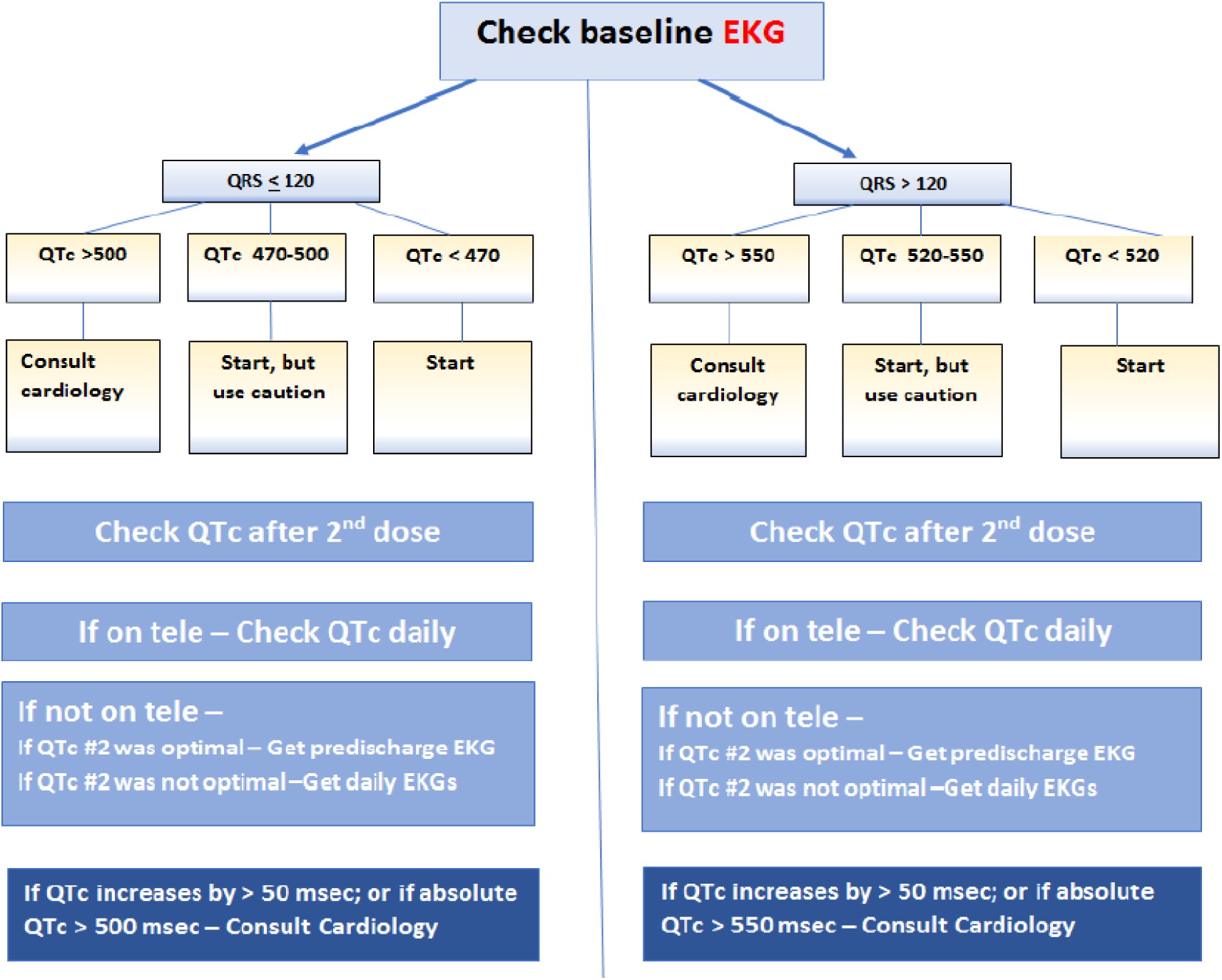
Cardiac Monitoring for patients on Hydroxychloroquine.

General Principles^61^
– **Given the growing evidence of myocarditis and arrhythmias with COVID, HCQ should be used with caution in this group of patients**.
– Obtain baseline EKG
– If on telemetry, check QTc and see if that corresponds to EKG QTc - If yes, use telemetry for further QTc monitoring. Otherwise Get EKG#2 and daily EKG as noted below.
– Discontinue all other QT prolonging agents
– Do not start Hydroxychloroquine if baseline QTc > 500 msec (or QTc > 550 msec in wide QRS patients) or discuss with cardiology if benefit vs risk is deemed high
– Be cautious if Baseline QTc > 470 msec (or QTc>520 msec in wide QRS patients)

- Check Telemetry QTc/ Acquire EKG#2 - preferably >2 hours after the 2nd dose of 400 mg Hydroxychloroquine.
- If QTc increases by less <50 msec; and if absolute QTc < 500 msec (<550 in wide QRS) - use lower dose.
- If QTc increases by >50 msec; or if absolute QTc > 500 msec (>550 in wide QRS) - use lower dose and recheck EKG daily for 2 days.
- Any evidence of Torsades on Tele -- D/c Hydroxychloroquine regardless of QT interval.
- Note - * Wide QRS defined as QRS > 120 msec.

